# Interprofessional collegiality and workplace abuse among healthcare workers in eastern Uganda: a mixed-methods study

**DOI:** 10.64898/2026.02.05.25342735

**Authors:** Ronald Tweheyo, Eric Oradi, Kennedy Pangholi, Olga Ariga Olaki, Barnabas Walozi, Kelvin Muwanguzi, Flavia Nakyejwe, Rhoda Mwesigwa, Joshua Epuitai

## Abstract

Interprofessional collegiality reflects mutual respect, empathy and solidarity within different health professions for a common goal. Limited studies have explored interprofessional collegiality in Uganda. This study aimed to determine interprofessional collegiality among healthcare workers in Eastern Uganda. We used a mixed-methods study design. The study was conducted among healthcare workers in two institutions in Eastern Uganda. We used the Practice Environment Scale to assess interprofessional collegiality among a sample size of 136 healthcare workers. The Likert Scale was used to assess the different dimensions of interprofessional collegiality. Ethical approval was obtained. Descriptive statistics were used for quantitative data, while thematic analysis was used for qualitative data. The majority of the participants were nurses/midwives (48.5%), allied health professionals (40.5%) and medical doctors (11%). The majority (65%) of participants strongly agreed and agreed that there was effective interprofessional collaboration in their workplace. Participants strongly agreed/agreed that there were good working relations (60%) and good teamwork (56%) between nurses/midwives and medical doctors. However, uncivil behaviours were common among healthcare workers, including psychological/emotional abuse (78%), physical abuse (4%), and sexual abuse (3%). In a qualitative study, uncivil behaviours occurred in the form of cold wars, name-calling, political interference, silent hatred, psychological stress, demotivation, absenteeism, and poor work relations. Poor collegial relations occurred from the individual (gender bias, perceived lack of capacity, poor supervision and leadership), interpersonal (lack of interpersonal respect, perceived lack of role clarity) and institutional factors (workload, poor working conditions, maldistribution of incentives). Despite the high interprofessional collegiality between nurses/midwives and medical doctors, workplace abuse among healthcare workers was high in our setting. Policymakers could prioritise strategies that address individual, interpersonal and institutional factors that result in poor work relations among healthcare workers.

## Introduction

Interprofessional collegiality (IPC) has been defined as *“a collective manifestation of respect, empathy and solidarity within the medical fraternity, driven by a common pursuit of clinical excellence in patient care, demonstrated through interpersonal/work relationships and organisational culture* (1)*.”* IPC represents a good working relationship, collaboration, and mutual support between two or more professional groups (1, 2).

IPC plays a key role in improving the quality of healthcare, patient safety, and patient outcomes, while it reduces healthcare expenditure (3). Good working relations enhance communication among team members, while they foster professional dignity and commitment to the profession (1, 2). Collegiality among healthcare workers can reduce conflicts in the healthcare setting and its associated consequences, such as emotional distress, poor communication, medical errors and poor patient outcomes (4–6). Uncivil professional behaviours are also linked to intentions to leave the healthcare profession, high attrition and turnover rates, and a shortage of the healthcare workforce (7, 8).

Uncivil professional behaviours are common among health workers globally (9). Uncivil behaviours hinder collegial mutual respect, empathy and solidarity among healthcare workers (1). The uncivil behaviours, referred to herein as workplace abuse, often occur in the form of bullying, mistreatment, and harassment, including the use of physical, sexual, emotional or psychological abuse(5). Evidence from a systematic review of 22 studies in Africa underscored widespread workplace abuse among healthcare workers, ranging from 9-100% (10). Similarly, in Uganda, the majority of healthcare workers experience verbal (64%) and physical abuse (32%) from their colleagues (11). In Greece, 50% of physicians did not fairly evaluate the nurses’ work, while 66% of them were insensitive to the needs of the nurses (12). The majority (60%) of nurses believed that physicians did not collaborate with them during therapeutic decision-making (12).

According to the Heise ecological model, uncivil behaviours are attributable to individual, interpersonal and institutional factors (13). Interprofessional conflicts and workplace abuse may stem from role conflict, poor communication skills, unrealistic expectations, and heavy workload (1, 14). Inter-professional conflicts emanate partly from lapses in leadership, remuneration structure, communication and emotional intelligence (15). In Uganda, unprofessional behaviours may occur from systemic challenges of low staffing, work absenteeism, occupational stress, work overload, low pay and poor working conditions(16, 17). Although in Uganda, good interprofessional relationships are valued, evidence on IPC and workplace abuse among health professionals remains limited (11). In Uganda, resource constraints may result in health systems deficiencies which ultimately affect collegiality and civil relationships among healthcare workers (19). Therefore, the study was conducted to determine the magnitude and drivers of interprofessional collegiality and workplace abuse among healthcare workers in Eastern Uganda.

## Methods

### Study design and setting

The study used a convergent parallel mixed-methods approach (20). The quantitative study was used to determine the magnitude of interprofessional collegiality and workplace abuse, while the qualitative study was used to understand the forms and drivers of workplace abuse. The findings were integrated into the discussion section (20). In Uganda, the healthcare system is based on a pyramidal referral model (18). Primary healthcare facilities form the base of the pyramid where they refer complicated cases to secondary and tertiary health facilities (18). The primary healthcare facilities comprise village health teams and health centers II and IIIs. Health center IVs and general hospitals are the secondary health facilities that refer to tertiary hospitals at the regional and national levels (18). Besides public health facilities, healthcare is also provided in the private for profit and private not for profit organizations under the public-private partnerships (18).

The study was conducted at a Regional Referral Hospital (RRH) and a public University in Eastern Uganda. The two sites were selected because they represented key teaching and healthcare institutions in eastern Uganda. The hospital was a high-volume facility with health workers of all cadres, including nurses, medical doctors, pharmacists and allied health professionals. In this study, clinical officers, anaesthetists, radiologists, laboratory technicians, and pharmacists have been referred to herein as allied health professionals. Healthcare workers in the hospital have different levels of training, which include certificate, diploma, bachelor’s and master’s levels. The university was chosen because it had diverse programs which attracted healthcare workers from various parts of the country, especially those who had come to acquire advanced qualifications. The university had healthcare workers with diploma qualifications, including nurses, clinical officers, midwives, laboratory personnel and healthcare workers with degree qualifications in medicine, nursing and pharmacy.

### Study Population and Sampling

This study included the health workers who were selected from the Regional Referral Hospital and a public Busitema university offering health professionals programs. The healthcare workers included nurses, midwives, doctors, pharmacists and allied health professionals. The sampling method used for quantitative data was convenience sampling, where participants were selected based on their availability at the time of data collection. The sample size was determined using the Kish-Leslie formula (21). The sample size was estimated based on the 90% magnitude of workplace abuse in Turkey (22). Consequently, a sample size of 136 participants was estimated in the study. For qualitative data, the purposive sampling method was used to collect diverse and in-depth data. We purposively selected participants with experience in uncivil professional behaviours, collegiality and workplace conflicts. The sample size was based on data saturation, which was considered when no new information or substantial insights were identified from the subsequent interviews (23). We reached saturation after the 13^th^ interview, while we conducted two additional interviews to confirm whether saturation was realised. This approach is consistent with the Hennink and Kaiser recommendation that saturation is often reached between 9 and 17 interviews (24).

### Study Variables

In this study, interprofessional collegiality was the dependent variable and it was measured using the Practice Environment Scale. The Practice Environment Scale involves five subscales that measure participation in hospital affairs, quality of care, support structures, staffing and resource adequacy and collegiality(25). The subscale on collegiality assesses teamwork, good working relationships and collaboration (joint practice) between nurses and physicians (25). The different responses were measured based on the Likert Scale categories of strongly agree, agree, neutral, disagree, and strongly disagree to the prepositions on the subscale (25). The Practice Environment Scale captures nurse-physician relationships. In our study, we adapted the tool to suit our setting and study population. Consequently, the nurse-physician dimension was expanded to include midwives and allied health professions. We modified the phrase physician in the scale to refer to doctors or allied health professionals. The modified adaptations were reviewed by two public health experts and piloted among 10 healthcare workers to ensure clarity and retention of the original construct validity. Secondly, we adopted the workplace abuse questionnaire to assess for uncivil behaviours (26). This included assessment of verbal abuse, physical abuse, sexual abuse or bullying of any form from the healthcare workers. Participants who experienced abuse were asked to identify the perpetrator of abuse. The questions on workplace abuse were adopted from another study (26).

### Data collection method

Data collection and recruitment of participants were done from March 15^th^ 2024, to August 15^th^ 2024. A self-administered questionnaire was used for data collection in the quantitative method. The questionnaire contained information on socio-demographics (age, sex, work experience, cadre, and level of education). The additional questions included position, title of job, marital status, communication skills, workload, and stereotypes. Interprofessional collegiality was assessed using the Practice Environment Scale (25). Data was collected in two different ways, both online using the Kobo toolbox and physically using printed questionnaires, depending on the comfort of the respondents.

In the qualitative study, in-depth interviews were carried out to enable a deeper understanding of the healthcare workers’ lived experiences regarding interprofessional collegiality and workplace abuse in eastern Uganda. An interviewer guide was developed by the research team, guided by the research objectives. Questions in the interview guide were open-ended, and they included questions on the levels of interprofessional collegiality, the perceived barriers and enablers to interprofessional collegiality and workplace violence/abuse, and specific experiences of verbal, physical, emotional or sexual abuse in the respective hospitals or health facilities. The in-depth interviews were conducted by RT and KP. Qualitative interviews were conducted in English in a safe and conducive environment. Interviews were audio recorded and lasted about 24 minutes to 48 minutes.

### Data analysis and management

Data was first entered into Microsoft Excel and later imported into STATA for analysis. Descriptive statistics, including frequencies and percentages, were used for categorical data and medians and interquartile range for continuous variables. For the qualitative arm, data were transcribed verbatim. Qualitative analysis was carried out using the Braun and Clarke thematic analysis (27). This involved reading through all the transcripts several times to gain familiarity with the data (27). Subsequently, initial codes, subthemes and themes were identified (27).

Qualitative data coding was performed independently by two researchers, and discrepancies were resolved unanimously. The generated codes were then grouped into themes where codes with similar meanings were fitted together into one theme (27). The themes formed were then reviewed and modified (27).

### Ethical approval

Ethical approval was obtained from the Mbale Regional Referral Hospital Research and Ethics Committee (MRRH-2023-356). Written informed consent was obtained from the participants. The ethical principles of research were maintained. Codes were used instead of the real names to protect the identity of participants.

### Trustworthiness and rigour of the study

The study used the Practice Environment Scale, which has remarkable validity and reliability (25). The modified questionnaire was pretested with 10 healthcare workers to assess the relevance, appropriateness, and feasibility of using the tool. In the qualitative study, measures were taken to address concerns of credibility, dependability, confirmability and transferability of the study findings (28). Credibility was addressed through interviewing different cadres of healthcare worker professionals, while bracketing and the use of two personnel to analyse the data may have addressed preconceived biases in the interpretation of the findings(28). We have used participants’ quotes to underscore the confirmability of our interpretations (28). We have described the study methods to enable replication and transferability of the study to other settings (28).

## Results

### Description of the participants

The median age of the participants was 27 years (interquartile range 25-32) (table 1). Out of the 136, 66 (48.5%) were either nurses or midwives, 15 (11.0%) were medical doctors, and 40 (55%) were allied health professionals (clinical officers, anaesthetists, radiologists, laboratory technicians, nutritionists and pharmacists). The majority103 (76%) of the participants described their work environment as being very good or good.

**Table 1:** Description of the participants.

| **Variable.** | **Frequency**<br>**(n=136)** | **Percentage** |
| --- | --- | --- |
| **Age**<br>median 27 (interquartile range: 25-32) |  |  |
| **Marital status**<br>Single<br>Married/cohabiting | 78<br>58 | 57.4<br>42.6 |
| **Sex**<br>Male<br>Female | 76<br>60 | 55.9<br>44.1 |
| **Level of education**<br>Certificate<br>Diploma<br>Bachelors/Masters | 26<br>52<br>58 | 19.1<br>38.2<br>42.7 |
| **Work experience**<br>Median (interquartile range) | 3 (1-7) |  |
| **Professional Background**<br>Nurse/midwife<br>Medical Doctor/specialist/consultant<br>Allied health professionals (clinical officer, anaesthetist, radiologist, lab technician, Pharmacists) | 66<br>15<br>55 | 48.5<br>11.0<br>40.5 |
| **The current job title matches the work experience/educational qualification.**<br>Yes<br>No | 123<br>13 | 90.4<br>9.6 |
| **Nature of work** |  |  |
| Supervisor/ward in charge/ manager | 32 | 23.5 |
| Under supervision | 104 | 76.5 |
| Number of people worked with (median and inter-quartile range) | 9 (5-16.5) |  |
| **Description of the work environment in the participant's workplace.** |  |  |
| Very good or good | 103 | 75.7 |
| Neutral | 30 | 22.1 |
| Poor | 3 | 2.2 |
| **Friend at the workplace (one participant socialises/ goes out with)** |  |  |
| Doctor | 36 | 26.5 |
| Nurse /midwife | 48 | 35.3 |
| Allied health professional | 33 | 24.2 |
| Most of them/all of them | 10 | 7.4 |
| None | 9 | 6.6 |
| **Staffing levels at the participant's workplace** |  |  |
| Adequate | 14 | 10.3 |
| Average | 102 | 75.0 |
| Poor or very low | 17 | 12.5 |
| Prefer not to say | 3 | 2.2 |

### Interprofessional collegiality

Concerning interprofessional collegiality, the majority of the participants strongly agreed/agreed that there was good collaboration 88 (65%), working relations 82 (60%), and teamwork 76 (56%) between nurses/midwives and medical doctors (table 2). In addition, the majority of the nurses/midwives strongly agreed/ agreed that they were understood 82 (60%) when they explained themselves. This was similar to those who agreed/strongly agreed to have been involved in policy-making or governance 82(60%), and that they felt supported even when in a direct conflict with a doctor 68(50%).

**Table 2:** Description of Interprofessional collegiality.

| **Variable** | **Frequency** | **Percentage** |
| --- | --- | --- |
|  | **n=136** |  |
| **The ward/hospital manager is supportive** |  |  |
| Agree/strongly agree | 100 | 73.5 |
| Neutral | 29 | 21.3 |
| Disagree/strongly disagree | 7 | 5.2 |
| **Senior midwives/ doctors/ nurses use mistakes as learning opportunities, not criticism.** |  |  |
| Agree/strongly agree. | 80 | 58.8 |
| Neutral | 32 | 23.5 |
| Disagree/strongly disagree. | 24 | 17.7 |
| **A midwife/doctor/nurse unit manager is a good manager and leader.** |  |  |
| Agree/strongly agree | 94 | 69.1 |
| Neutral | 31 | 22.8 |
| Disagree/strongly disagree | 11 | 8.1 |
| **Midwife/doctor/ nurse praises and recognises others for a job well done.** |  |  |
| Agree/strongly agree. | 83 | 61.0 |
| Neutral | 35 | 26.0 |
| Disagree/strongly disagree. | 18 | 13.0 |
| **A midwife/doctor/nurse unit manager backs up the staff in decision-making, even if the conflict is with a doctor.** |  |  |
| Agree/strongly agree | 68 | 50.0 |
| Neutral | 41 | 30.1 |
| Disagree/strongly disagree | 27 | 19.9 |
| **Career development/clinical ladder.** |  |  |
| Agree/strongly agree. | 79 | 58.1 |
| Neutral | 41 | 30.2 |
| Disagree/strongly disagree. | 16 | 11.7 |
| **Opportunity for midwives to participate in policy decision-making governance.** |  |  |
| Agree/strongly agree | 81 | 59.6 |
| Neutral | 35 | 25.7 |
| Disagree/strongly disagree | 20 | 14.7 |
| **Hospital managers consult about daily problems.** |  |  |
| Agree/strongly agree. | 76 | 55.9 |
| Neutral | 35 | 25.7 |
| Disagree/strongly disagree. | 25 | 18.4 |
| **Midwives feel understood when they explain themselves.** |  |  |
| Agree/strongly agree | 82 | 60.3 |
| Neutral | 34 | 25.0 |
| Disagree/strongly disagree | 20 | 14.7 |
| **Enough healthcare workers to provide quality patient care in my facility.** |  |  |
| Agree/strongly agree | 53 | 38.2 |
| Neutral | 33 | 24.3 |
| Disagree/strongly disagree | 51 | 37.5 |
| **Enough staff to get the work done in my facility.** |  |  |
| Agree/strongly agree | 50 | 36.8 |
| Neutral | 43 | 31.6 |
| Disagree/strongly disagree | 43 | 31.6 |
| **High standards of health care are expected by the hospital management.** |  |  |
| Agree/strongly agree. | 90 | 66.2 |
| Neutral | 29 | 21.3 |
| Disagree/strongly disagree. | 17 | 12.5 |
| **Doctors and midwives/nurses have good working relations.** |  |  |
| Agree/strongly agree | 82 | 60.3 |
| Neutral | 36 | 26.5 |
| Disagree/strongly disagree | 18 | 13.2 |
| **There is good teamwork between midwives/nurses and doctors.** |  |  |
| Agree/strongly agree | 76 | 55.9 |
| Neutral | 42 | 30.9 |
| Disagree/strongly disagree | 18 | 13.2 |
| **There is collaboration (joint practice) between midwives/nurses and doctors.** |  |  |
| Agree/strongly agree | 88 | 64.7 |
| Neutral | 35 | 25.7 |
| Disagree/strongly disagree | 13 | 9.6 |

### Work-related abuse among healthcare professionals

More than two-thirds 106, 77.9%) of the healthcare workers had at least experienced one form of psychological/ emotional abuse from other healthcare professionals (table 3). The emotional/psychological abuse was in the form of belittling/undermining 47 (34%), persistent humiliation in front of others 43(32%), intimidation 36(26%), and teasing 32(24%), among other forms of abuse.

**Table 3:**
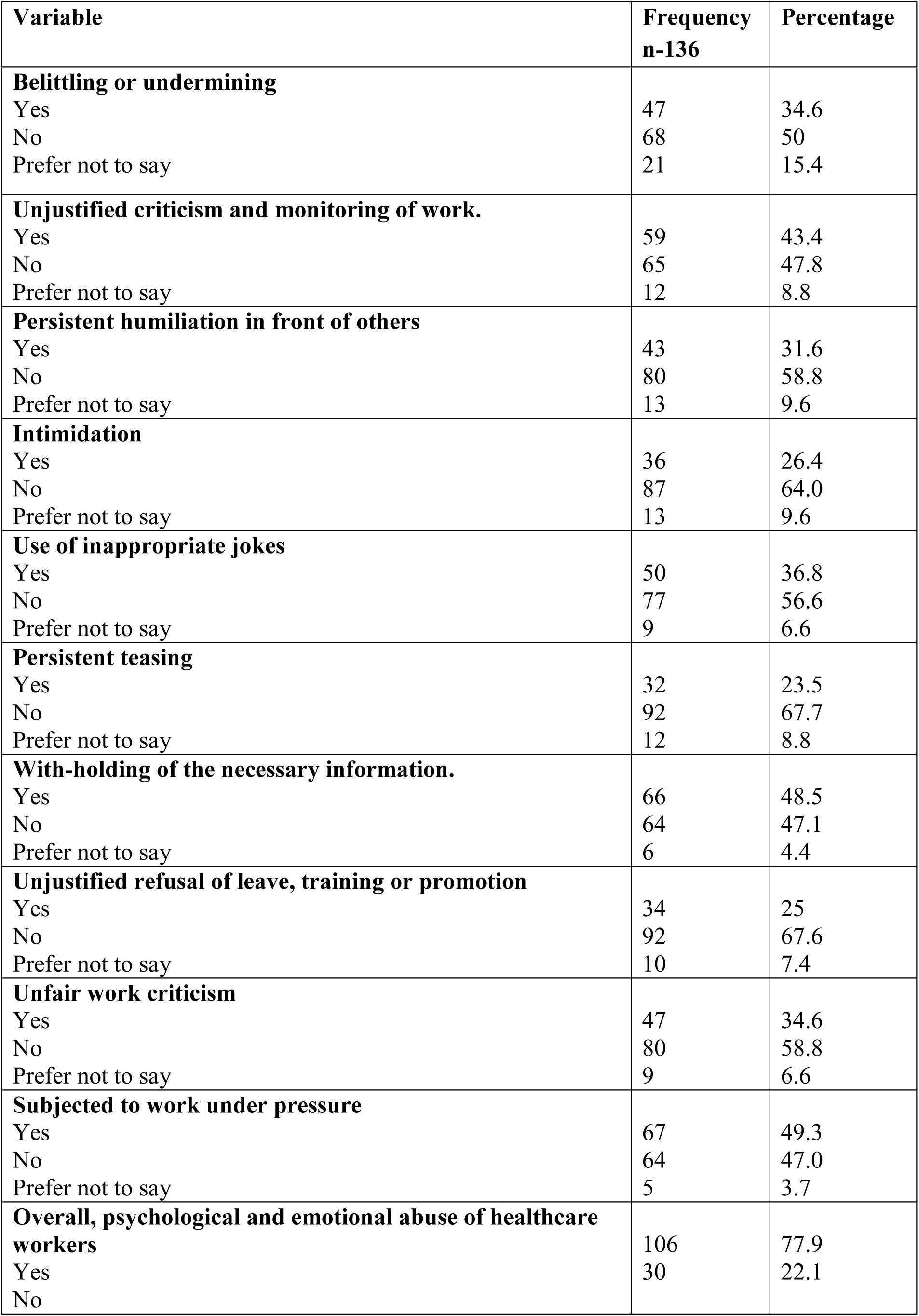
Psychological and emotional abuse of healthcare workers multiple responses.

### Prevalence of sexual and physical abuse by healthcare professionals

Out of the 136 healthcare workers, 3.7 % experienced sexual abuse, while 4% experienced physical abuse from fellow healthcare professionals (**Error! Reference source not found.**).

**Figure 1:**
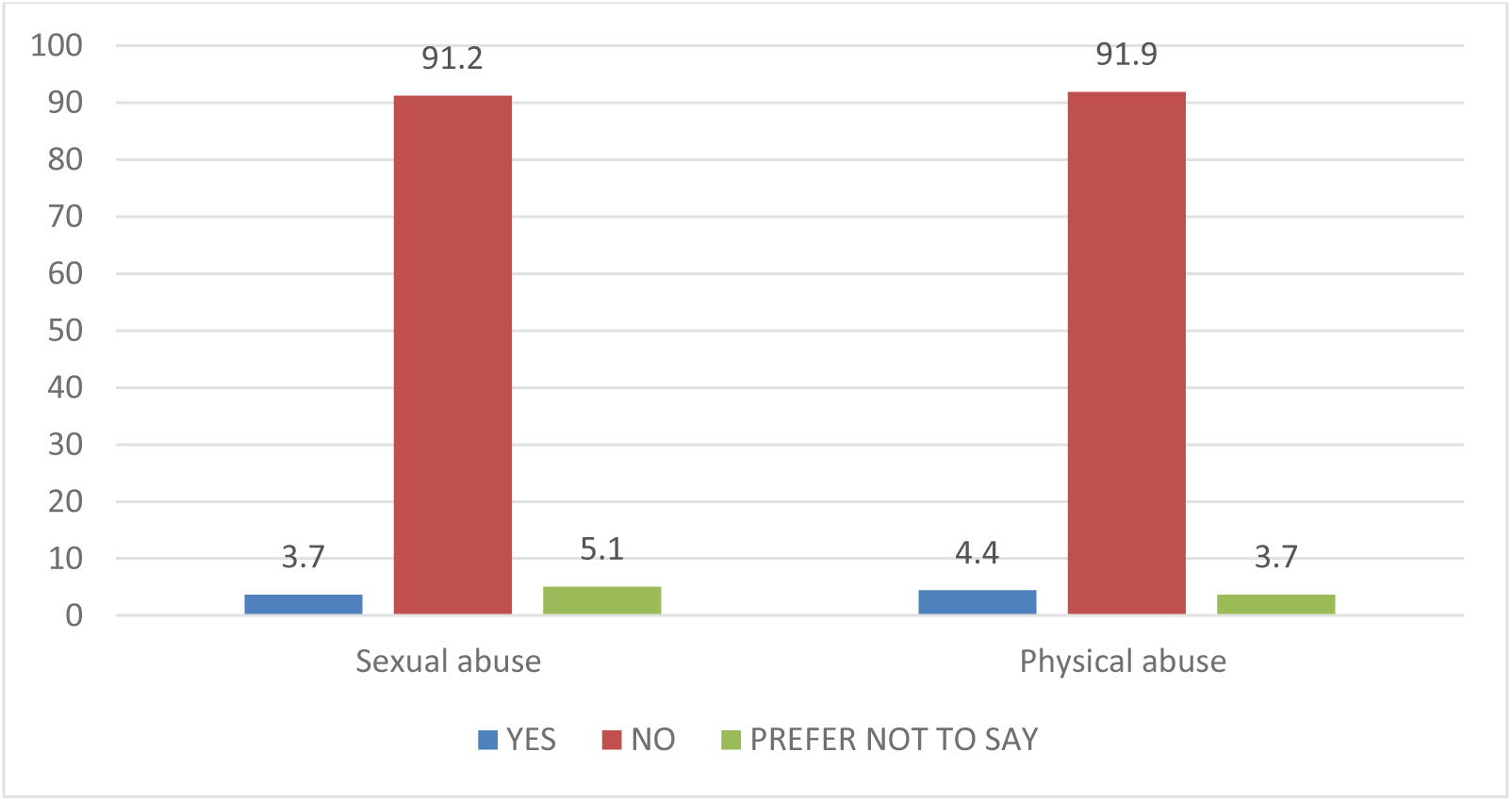
Experience of sexual and physical abuse among healthcare workers.

### Perpetrators of abuse against healthcare workers

Overall, the main perpetrators of abuse were nurses/midwives (46.5%), followed by allied health (25.6%) and medical doctors (24.4 %), while 3.5% of participants reported having been harassed by all professions (figure 2)

**Figure 2:**
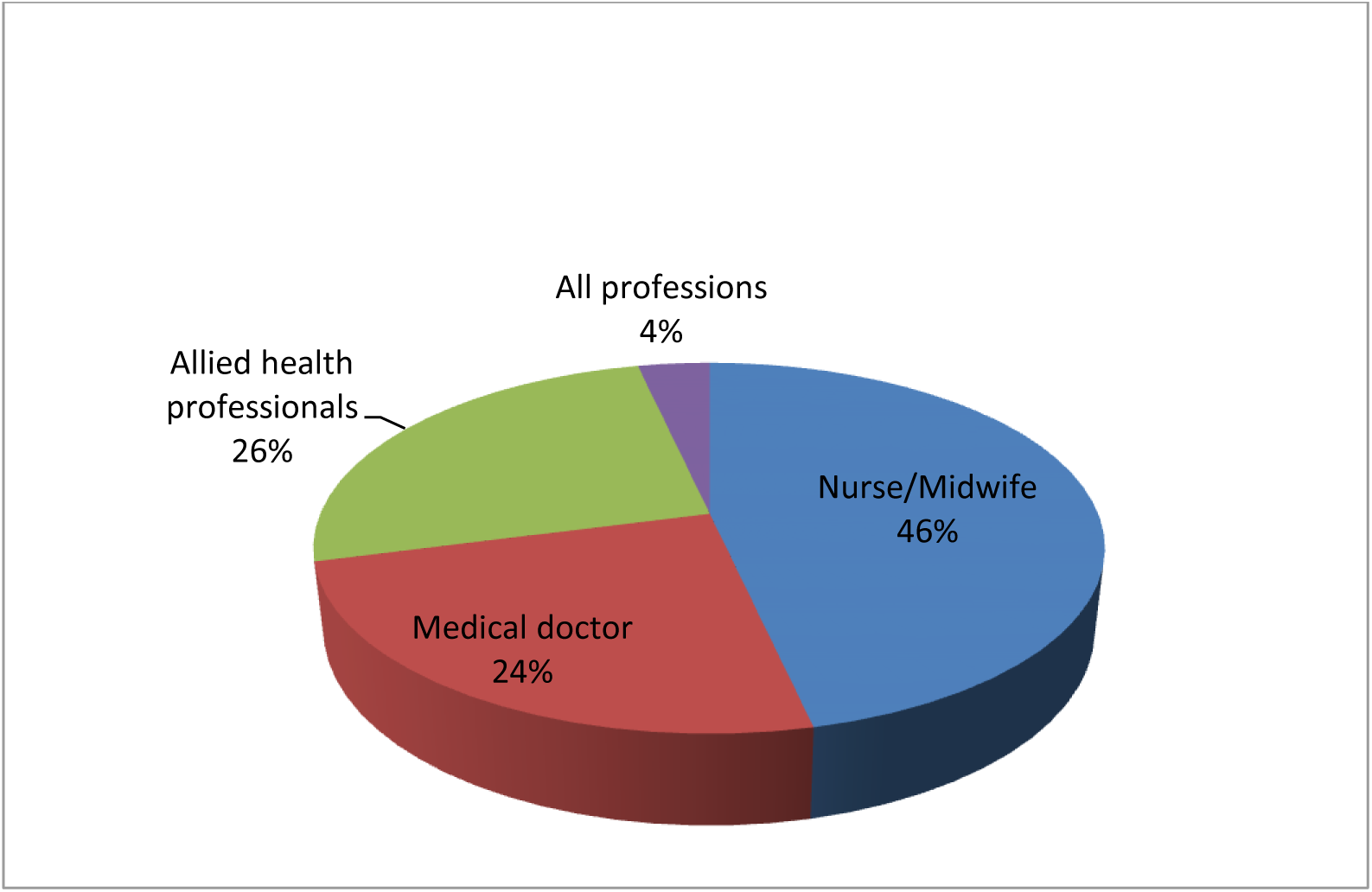
Perpetrators of emotional, physical and sexual abuse against health workers.

### Qualitative data

In the qualitative study, we conducted 15 in-depth interviews with 10 males (66.7%) and five females (33.3%) (Table 4). In this study, nurses 4(26.7%) and clinical officers 11 (73.3%) were recruited. Pseudonyms such as R1, R2, and R3 were assigned to each participant to conceal their identities.

**Table 4:** Description of the study participants.

| Variable | Frequency | Percentage |
| --- | --- | --- |
| **Age** |  |  |
| 25-35 | 13 | 86.7 |
| 35+ | 2 | 13.3 |
| **Sex** |  |  |
| Male | 10 | 66.7 |
| Female | 5 | 33.3 |
| **Profession** |  |  |
| Nurse | 4 | 26.7 |
| Clinical officer | 11 | 73.3 |
| **Marital status** |  |  |
| Married /cohabiting | 11 | 73.3 |
| Single | 4 | 26.7 |
| **Education level** |  |  |
| Diploma | 15 | 100% |

### Theme 1: Forms of mistreatment and its consequences

Participants identified experiencing various forms of abuse, such as psychological and emotional abuse, verbal abuse, and sexual and physical abuse. Other additional concerns included political interference and denial of training opportunities.

### Subtheme: Psychological and emotional abuse

Emotional abuse occurred in the form of cold wars and name-calling. Healthcare workers employed cold war tactics such as avoiding each other, silent hatred, and work absenteeism:

> *"You see people having their beef, but they don’t speak, but it is in the form of withdrawals from work…..I can mean to withdraw from performing my duties." (R6, clinical officer who is in charge).*

Name-calling was usually reserved for supervisors or colleagues as a form of torture for colleagues at the workplace.

> *"That one is very common nick-naming. It happens as in the juniors nicknaming the supervisors or the bosses. Someone can call you a policeman, like you hear them saying a policeman has come." (R7, a senior clinical officer in a government institutional facility)*.

### Subtheme: Verbal abuse

Healthcare workers reported experiencing verbal abuse, which was in the form of backbiting, abuse, and shouting at others. Verbal abuse involved health workers talking negatively about their colleagues in their absence. In some cases, overt verbal abuse included hurling insults, abuse, and shouting at colleagues. Verbal abuse often occurred between the supervisor and the subordinates.

> *"I saw a senior surgeon still in the theatre [with] an intern doctor…during an operation, and he became too tough on her. He used a lot of bad words, and the lady cried, and she even failed to participate in the surgery.*” (R13 an assistant nursing officer).

### Subtheme: Physical abuse

Some of the healthcare workers experienced physical abuse from their colleagues. The physical abuse included being beaten or slapped. Physical abuse occurred from healthcare workers and sometimes from employers who were not medical personnel.

> *"…she wanted me to change the prescription, I thought that no, I’m not changing it…She slapped me."* (R11, a clinical officer in a surgical hospital).

### Subtheme: Sexual harassment

Sexual harassment was thought to be more common in the workplace. This was perceived to usually occur between bosses and subordinates, between seniors and juniors, and between the young and the very old.

> *"I have seen a lot of sexual violence among health workers, especially between juniors and seniors, even those with a very big age difference, because of greed and wanting to use the authority they have over others." (*R5, a clinical officer).

### Subtheme: Other forms of abuse and conflicts

Political interference was another form of abuse that health workers experienced during work. Healthcare workers who were thought to have connections to the political wing refused to work. The supervisors feared being negatively reported to politicians for fear of losing their jobs.

> *" So, the nurse of this lower carder sometimes they don’t want to do things because they know they will tell the supervisor like that political interference.”(*R9, a senior clinical officer in charge of Health Centre III).

Some health workers reported how unfairly they were denied participation in training, and yet they believed that they were entitled to them. They further cited that those who went for training would not arrange educational sessions for their colleagues, but they would instead wait to scold them on how they were knowledge-deficient.

> “*Imagine, like every training, you’re the one attending. But when you come back, you do not want to put a CME [continuous medical education] for the rest to also learn from what you learned, but you’re waiting for them to make a mistake and then you tell them no, you do not know …there’s nothing you know like that."(*R10, a Female clinical officer)

### Subtheme: Response to abuse and mistreatment

Work-related conflicts and abuse were perceived to lead to poor patient outcomes, demotivation and psychological trauma. The psychological effects included stress and depression, while demotivation resulted in abscondment from work, employee turnover and leaving the job.

Healthcare workers who experienced conflicts displaced their anger toward patients. Some preferred to use bureaucratic processes to delay patient care. This was common when healthcare workers had conflicts with a person who was caring for the respective patient.

> *"If the person in the clinical room has beef [conflict] with the person in the lab, when this one sends lab requests, the other may ignore them, or it will take long to produce results for the patient, affecting the patient"* (R 5, a clinical officer who works in an HIV unit).

Poor interprofessional relationships were seen to deter others from getting promotions at work. In some cases, healthcare workers were denied training opportunities and incentives because of workplace abuse.

> *"If you have an opportunity like a training…and you see this person is not taken, that is a little bit of psychological torture, and yet it is in their area of jurisdiction….but they are not taken because they are not in a good relationship with the bosses. It kind of like torments them.”* (R7, a clinical officer, a senior staff).

### Theme 2: Drivers of interpersonal conflicts among health workers

Participants identified various drivers of the mistreatment of healthcare workers. These included individual (perceived lack of capacity, leadership and supervision, gender bias), interpersonal (lack of respect for other professions) and institutional factors (maldistribution of incentives, workload and working conditions, poor communication).

### Subtheme: Perceived lack of capacity

Interpersonal conflicts among healthcare workers were driven by a perceived lack of capacity, particularly related to discrepancies in the supervision hierarchies. While a person of a higher qualification and work experience would usually be the one to supervise someone with lower qualifications or experience, there were instances when subordinates supervised a person with higher qualifications. Conflicts were thought to arise when the less qualified supervised those with higher qualifications. Conflicts also occurred when the young were given to supervise older healthcare providers.

> *"A health centre three supposed to be headed by someone of diploma…, so you find someone of a certificate in charge of that facility and they leave out someone with a diploma because of politics."* (R6, a senior clinical officer).

Besides, interprofessional conflicts due to the supervisory role, lower level of education than the subordinates or having a supervisor with the same level of education as the subordinates were also perceived to cause work conflicts. This was common in local government, where clinical officers with diplomas were made to head health facilities. Nurses who had a diploma qualification, like the clinical officers, did not take it well, which resulted in ongoing conflicts. In addition, conflicts also occurred when supervisors had the same salary scale as the subordinates.

> *"A clinical officer heading a health centre is at the same education level as a diploma nurse who will not take it well, and mostly it creates kind of like a conflict, and usually if you don’t have a good working relationship, they will let you make mistakes…they cannot correct you, but they do finger-pointing."* (R9, Senior clinical officer in charge of a health centre III)

### Subtheme: Gender bias

Gender bias and inequalities exacerbated social-related abuse in the workplace. Work conflicts were common when females were given managerial or supervisory roles. Conflicts were seen to occur when females were supervising both male and female colleagues. Female supervisors were thought to disrespect and disregard the needs and demands of their female colleagues. This reflects broader systemic gender biases and challenges in leadership in hierarchical systems, which directly resulted in workplace abuse of health workers.

> *"Working with a manager who is female, and you are also female, I don’t know if it’s biased, but they always have kind of disrespect for their fellow females, ordering them around, not respecting their decisions, they are not willing to listen to them and being too bossy to them."* (R13, a female assistant nursing officer at a government facility).

### Subtheme: Poor supervision and leadership skills

Poor supervision and leadership skills were perceived to cause interprofessional conflicts among healthcare providers. Lack of exemplary leadership was seen to result in absenteeism among subordinates. Supervisors who delegated work to subordinates but with limited power and decision-making were seen to demotivate and cause work-related conflicts. In some cases, supervisors could delegate work, but they were reluctant for the subordinate to get the associated benefits, such as monetary incentives.

> *"Supervisor is delegating you to stand in for them, but then they give you so limited powers to do some of the things. They will tell you not to do it until they return, just because maybe there is a money/financial implication in them"* (R7, a senior clinical officer).

While there was a challenge of poor leadership and supervisory roles, having visionary leadership with a sense of direction and good monitoring minimised conflicts at work. Adequate and proper monitoring of health workers was seen to minimise conflicts at work.

> *“If people knew that they were being monitored, they would also keep within their lane and these sources of conflicts would not be coming” (*R9, a senior clinical officer in charge of Health Centre III.)

Gaps in leadership manifested in the form of a lack of communication or a lack of regular meetings among staff. The lack of meetings inhibited the resolution of conflicts and uneasiness among healthcare providers. This was seen to result in toxic work environments and work-related conflicts.

> *"If you don’t communicate well, if you don’t have regular meetings with staff and are not happy about something, they tell you that you change so that you create a harmonious environment, but if such kind of things don’t go on, conflicts happen."* (R7, a senior clinical officer)

### Subtheme: Lack of respect for other professions

Lack of respect among healthcare providers was seen to cause work-related conflicts. Lack of respect was brought about by personality differences, lack of respect for other professions, and differential access to opportunities and resources. Some healthcare providers perceived themselves to be superior, more knowledgeable and infallible. This failed to acknowledge their limitations, weaknesses and knowledge gaps, but it also led to a lack of collegial relations among healthcare providers.

> *"Most fights between health workers are always about who knows more and who knows less. Any cadre is always beefing the other that for us we know much and for you, you know less, like surgeons are beefing physicians and physicians are also doing the same."* (R3, a clinical officer formerly working in a research organization).

> “*The health workers, like doctors, who will feel that they are so superior above everyone else, and they cannot be corrected…Just because of your ego, and you feel like I’m a doctor, I cannot listen. I am not corrected by anyone below me. Such kinds of things cause a lot of conflicts at work."* (R10, a Female clinical officer.).

Lack of respect for other professions also occurred because of a lack of role clarity and the blurring of roles. The blurring of roles occurred due to task shifting amidst shortages of skilled healthcare providers.

> *“Lack of role clarity contributes a lot to conflicts. You realize that they do what they are not supposed to do and at times they do it wrongly.”* (R6, a senior clinical officer.)

### Subtheme: Maldistribution of resources and incentives

Most of the conflicts among healthcare workers arose from the maldistribution of resources, training opportunities, and work-related incentives. Supervisors often misappropriated and maldistributed incentives meant for their staff for personal gain. As such, healthcare providers did not get their incentives and allowances, which resulted in bickering, conflicts and demotivation for work. Besides the misdistribution of incentives, work-related conflicts were attributed to denying training opportunities to the subordinates. Training opportunities, besides acquiring knowledge and skills, attracted significant financial gains. As a result, conflicts occurred when the supervisor or a particular group of personnel were often selected for training while others were being ignored.

*"Like if there’s a training and one is supposed to be in that training….but your supervisor will not allow you to go for that training because they don’t want you to …go… in case it’s mostly related to financial gains"* (R7, a male senior clinical officer)

### Subtheme: Work overload and poor working conditions

The fights were also due to work overload. The excess workload was thought to cause stress and burnout, leading to more conflicts. While work overload was due to a shortage of staff, some health workers intentionally refused to work. Absenteeism of some healthcare workers was seen to annoy others, resulting in work-related conflicts. Participants also revealed that low salary and a poor working environment were major drivers of workplace violence among health workers.

> *"Many people out there point a lot of figures to midwives, but I think the kind of workload that midwives have has subjected them to stress. A midwife has been placed in a health Centre III, which is having too much work on a night shift alone, so you come when you have burnout*.” (R2, a male assistant nursing officer at a health Centre.)

Health workers sharing houses or a lack of proper accommodation resulted in conflicts among healthcare workers who were staying together.

> *"There is no better housing… people have been forced to share houses. and that brings in grudges between the two people." (R10, a female clinical officer)*.

### Subtheme: Poor communication skills

Poor communication skills and the use of abusive or harsh language were seen to foster workplace abuse among healthcare workers. The use of kind and respectful language, especially among healthcare workers in administrative or managerial positions, was seen to promote collegial relations among healthcare workers.

> *“Doctors interacting with the lower cadres, they are humble, they respect them” (*R13, an assistant nursing officer at a government facility).

## Discussion

The study was conducted to determine the level of interprofessional collegiality and workplace abuse among healthcare workers. Overall interprofessional collegiality, which was measured in terms of work relations, collaboration, and teamwork, was noted to be good between medical doctors and nurses/midwives. However, workplace abuse was also reported at high rates, with over 78% of healthcare workers experiencing at least one form of psychological abuse. Physical and sexual abuse was reported among 4% of healthcare workers. In the qualitative interviews, diverse forms of workplace abuse were noted, including political interference, name-calling, cold wars, and verbal abuse. The drivers of conflicts were related to perceived lack of capacity, gender bias, role clarity, leadership and institutional factors (e.g., work overload and maldistribution of resources). Our study findings have important implications regarding collegiality and work dynamics among healthcare professionals.

In this study, interprofessional collegiality between midwives/nurses and doctors was high. This was similar to a study in West Ethiopia, where 67% of the participants had a satisfactory interprofessional collaboration between nurses and physicians (29) and the 73% ideal teamwork observed in Slovakia (30). Although interprofessional collegiality was reported to be high in our setting, the majority (78%) of healthcare workers experienced workplace abuse. The magnitude of abuse amidst the high collegiality noted in our study may reflect superficial collaborations and teamwork, while strained interprofessional relationships exist beneath the tip of the iceberg.

Normalisation of workplace abuse, hesitancy to report experience of abuse and social desirability bias could explain the high interprofessional collegiality in our study despite experiences of widespread abuse(31).

The drivers of workplace abuse stem from individual, interpersonal and institutional factors(14). While studies have identified female healthcare workers as the main victims of workplace abuse, including sexual harassment (32), our study highlighted gender bias as one of the drivers of workplace abuse. Conflicts seemed to stem among female healthcare workers, especially when they were in positions of leadership. The gender bias highlights the wider societal gender inequalities and stereotypes that seem to pervade the healthcare system, fostering workplace abuse (14, 33). This calls for the need to train healthcare workers on culturally appropriate interventions of conflict resolution, while emphasising the importance of cultural and gender-sensitivity, role clarity, interprofessional collaboration, respect and collegiality.

Maldistribution of resources, incentives, and promotions was seen to promote workplace abuse. Individuals in lower positions may occasionally be given the responsibility to supervise those in senior positions, which can promote workplace abuse from perceived incompetence of the supervising healthcare workers. Inequities in resource allocation, especially sharing incentives, resulted in demotivation, lack of empowerment and workplace abuse among healthcare workers.

Furthermore, workplace abuse was seen to occur in the setting of work overload, poor working conditions and poor accommodation services for healthcare workers(34, 35). In Low and middle-income countries such as Uganda, health system challenges, including staffing, remuneration, leadership and governance, and work conditions, may have further exacerbated the poor work relations(36). Addressing institutional factors and strengthening leadership and support supervision may play a role in reducing workplace abuse among healthcare workers (37).

Workplace abuse results in negative social, psychological, cognitive and emotional consequences(1). In our study, responses to workplace abuse varied from demotivation, sabotage, and disengagement to work absenteeism and abscondment. Individuals who had conflicts at work used cold war tactics and bureaucratic strategies to sabotage their colleagues’ work. These forms of interprofessional hostility compromise both staff well-being and patient care (1). Consistent with previous studies (35), healthcare workers had emotional responses to abuse, including work-related stress and depression (38). This underscores the negative impact of workplace abuse on the mental health of healthcare workers (32). Workplace abuse amidst mental health struggles may lead to high turnover and attrition rates among healthcare workers, as individuals may develop intentions to leave the workplace(7, 8). High health workforce turnover and attrition rates will compound the shortage of healthcare workers and the high healthcare provider-to-patient ratio. This underscores the importance of addressing the consequences of workplace abuse (36).

### Study strengths and limitations

The study used mixed methods to understand the phenomenon of collegiality and hostility in the workplace. We did not determine the factors associated with collegiality in our setting. Nevertheless, qualitative findings in our study were used to provide rich contextual understanding of the factors which were perceived to be associated with collegiality and hostility in the workplace. The sensitivity of abuse, recall bias, and social desirability bias may have resulted in hesitancy to report experiences of abuse and the likely tendency to report high interprofessional collaboration (39). The qualitative study relied mostly on nurses and allied health professionals. As a result, our findings may not be transferable to medical doctors. The study findings may not be transferable to dissimilar settings. In health care settings, power dynamics between the interviewers and participants, especially in hierarchical settings, might have influenced the quality of the interviews.

## Conclusion

The majority of the participants agreed that there was effective interprofessional collaboration (65%), good working relations (60%), and good teamwork (56%) between nurses/midwives and medical doctors. Ultimately, our study noted good interprofessional collegiality among healthcare workers. However, 78% of the health workers experienced at least one form of psychological or emotional abuse, while 4% of healthcare workers experienced physical abuse, and 4% of them experienced sexual violence during work. Uncivil behaviours emanated from disharmony with supervision hierarchies, gaps in leadership and supervision, gender bias, inequitable distribution of incentives, lack of respect for other professions, and work overload. Addressing the drivers of uncivil behaviours is needed to promote interprofessional collegiality. Policies need to be developed to promote collegial interactions among healthcare providers.

Enforcement of these policies and guidelines could help ensure safety and respectful workplaces in healthcare settings. This can occur through strengthening mechanisms of reporting of unprofessional vices through use of anonymous online reporting mechanisms. Future studies could be conducted among medical doctors to explore their experiences of workplace abuse.

## Data Availability

All data produced in the present study are available upon reasonable request to the authors

## Abbreviations

IPC: Interprofessional collegiality/ Interprofessional collaboration
WHO: World Health Organisation
MRRH: Mbale Regional Referral Hospital
WPV: Workplace Violence
NGO: Non-Governmental Organisation
UK: United Kingdom
USA: United States of America
US: United States
SD: Standard Deviation
BUFHS: Busitema University Faculty of Health Sciences

## Declaration of Competing Interest

The authors declare no competing interests.

## Availability of data and materials

Additional data and materials can be accessed on request from the corresponding author.

## Funding

The research was not funded.

## Author’s statement

TR, JE &OE were involved in the conceptualisation of the study. JE and TR designed the study; OE, WB, MK, PK, NF, MR and OAO collected data. JE and TR analysed the data and drafted the manuscript. JE reviewed the manuscript. JE provided the overall oversight in the design and implementation of the study. All authors meet the criteria for author contributions and have read and approved the manuscript.

## Acknowledgements

We appreciate the participants who agreed to take part in this study.

## Notes

### Competing Interest Statement

The authors have declared no competing interest.

### Funding Statement

This study did not receive any funding

